# AI for Early Warning of Seasonal Infectious Disease: Shapely Additive Explanations Improves Prediction of Extraordinary West Nile virus Events in Europe

**DOI:** 10.1101/2020.08.27.20183327

**Authors:** Albert A Gayle

## Abstract

West Nile virus disease is a growing issue with devastating outbreaks and linkage to climate. It’s a complex disease with many factors contributing to emergence and spread. High-performance machine learning models, such as XGBoost, hold potential for development of predictive models which performs well with complex diseases like West Nile virus disease. Such models furthermore allow for expanded ability to discover biological, ecological, social and clinical associations as well as interaction effects. In 1951, a deductive method based on cooperative game theory was introduced: Shapley values. The Shapley method has since been shown to be the only way to derive “true” effect estimations from complex systems. Up till recently, however, wide-scale application has been computationally prohibitive. Herein, we present a novel implementation of the Shapley method applied to machine learning to derive high-quality effect estimations. We set out to apply this method to study the drivers of and predict West Nile virus in Europe. Model validity was furthermore tested using observed information in the time periods following the prospective prediction window. We furthermore benchmarked results of XGBoost models against equivalently specified logistic regression models. High predictive performance was consistently observed. All models were statistically equivalent in terms of AUC performance (96.3% average). The top features across models were found to be vapor pressure, the autoregressive past year’s feature, maximum temperature, wind speed, and local GNP. Moreover, when aggregated across quarters, we found that the effect of these features are broadly consistent across model configurations. We furthermore confirmed that for an equivalent level of model sophistication, XGBoost and logistic regressions performed similarly, with an advantage to XGBoost as model complexity increased. Our findings highlight the importance of ecological factors, such as climate, in determining outbreak risk of West Nile virus in Europe. We conclude by demonstrating the feasibility of same-year prospective early warning models that combine same-year observed climate with autoregressive geospatial covariates and long-term bioclimatic features. Scenario-based forecasts could likely be developed using similar methods, to provide for long-term intervention and resource planning, therefore increasing public health preparedness and resilience.

Highlights
- For geospatial analysis, XGBoost’s high-powered predictions are not always empirically sound
- SHAP, an AI-driven enhancement to XGBoost, resolves this issue by: 1) deriving empirically-valid models for each individual case-region, and 2) setting classification thresholds accordingly
- SHAP therefore allows for predictive consistency across models and improved generalizeability
- Aggregate effect estimations produced by SHAP are consistent across model configurations
- AI-driven methods improve model validity with respect to predicted range and determinants

## Background

West Nile Virus (WNV) is a mosquito-borne virus with deadly potential^1^. The virus has since rapidly spread in recent years and now enjoys the widest geographic distribution of any similar mosquito-borne viruse^2^. 2018 alone saw a 7.2 fold increase in the number of cases reported in Europe along with a markedly expanded geographic range (see *Figure 1*). Many factors have been associated with this spread, but much is still unknown^3^. The endemic potential of WNV is not to be underestimated: seropositivity is reportedly as high as 90% in the sub-Saharan regions where WNV has long been endemic^4^. Reports of human-to-human transmission routes – via blood transfusion^5^, organ transplants^6^, aerosol^7,8^, birthing and breast feeding^9^, and possibly even sexual contact^10^ – have added to the concern^11,12^. Response options are limited to symptomatic treatment and no human vaccine is available^10^. And the continued emergence of multiple lineages^13–16^, each with distinct transmission and syndromatic profiles^17^, adds to the uncertainties. Predicting annual emergence and spread of WNV in Europe has therefore become a priority.

**Figure 1a,b,c.**
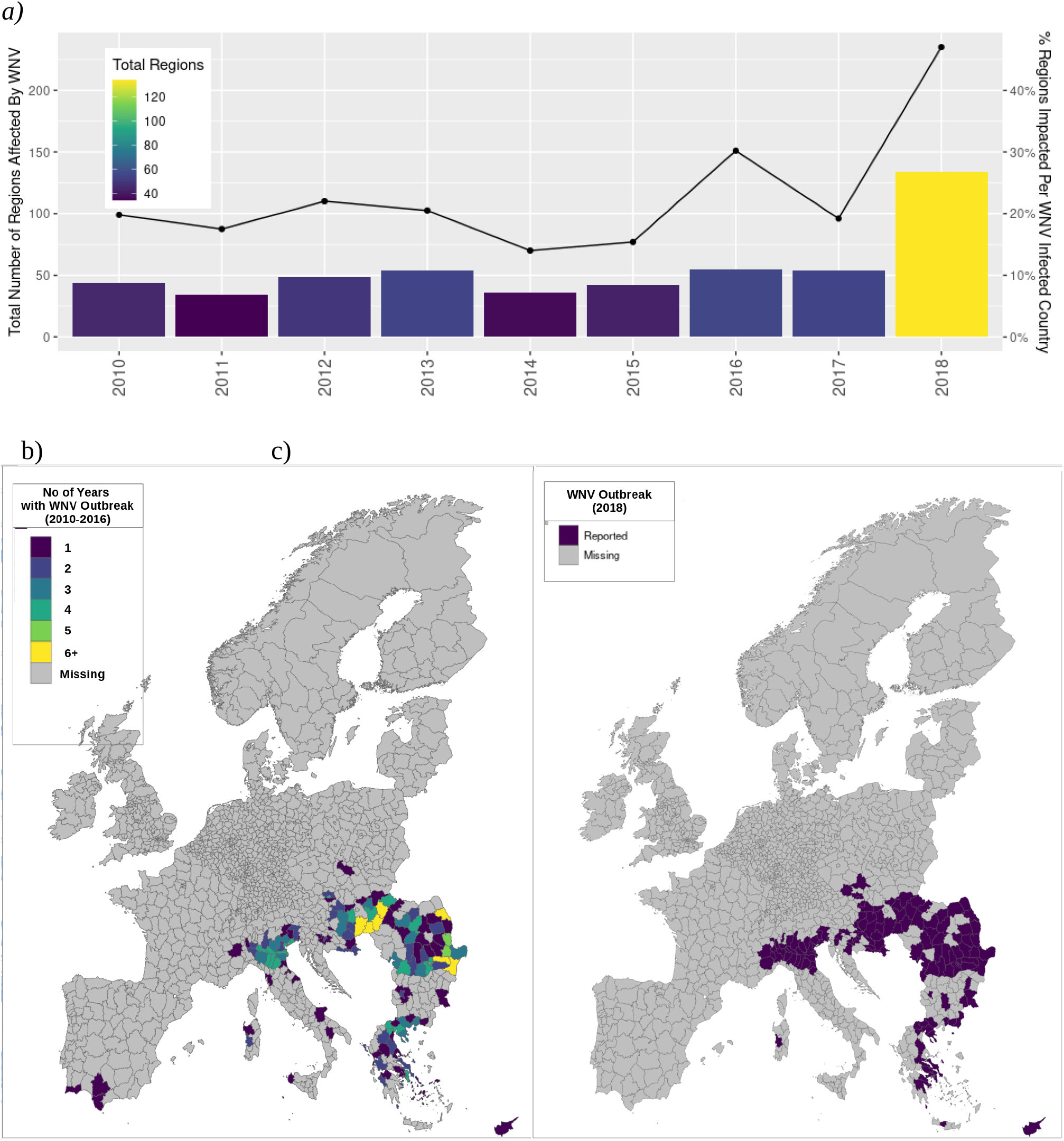
Spatiotemporal distribution of WNV infection and ecoclimatic trends. Our study included 1811 NUTS3 subregions comprising 29 countries within the EU/EEA (*Supplementary Table 1*). During the study period, WNV outbreaks were found to occur sporadically but with an overall increasing trend in terms of number of WNV cases reported per affected country per year and number of regions reporting WNV cases per affected country per year. This trend was driven primarily by the extraordinary 2018 outbreak year in which more cases were reported than in the preceding 7 years combined. *(Figure 1a)*. The geospatial spread of WNV infection during the 2018 event year (*Figure 1b)* is broadly equivalent to the cumulative range from the 2010-2016 control period *(Figure 1c)*.

The spread of West Nile virus is dependent on a wide range of determinants, including climatic features, environment, and sociodemographic factors^18^. However, predictive modeling efforts have often been confounded by complex interactions and time-varying associations^19^. Early warning models can therefore often be difficult to implement in practice. Various analytical modalities have been used for this purpose, each with different pros and cons. Most have met with varying degrees of success, but scope- or context-dependent inconsistency of effect remains an outstanding issue. Classification tree models are a promising entry in this field. However, some controversy has been merited. Most notably, questions persist as to the long-term predictive accuracy of these models. This is due to the “black box” nature of these models whereby the mechanistic structure being assumed by these models are not open to scrutiny: unlike traditional models, no parameter estimates are provided. This makes such models particularly unsuited for the early warning case, or intervention strategy, as 1) direct effect of specific predictors on the predictive outcomes cannot be determined, and 2) there is no way to control or investigate the reliability of the deterministric structure driving predictions.

In this paper, we demonstrate application of a novel AI-driven solution with potential to overcome the problems associated with classification tree models. The novel SHAP (SHaply Additive Explanation) framework^20^ deductively imputes feature effects for each individual case^21^. (Refer to *Methods* for further details.) The resulting meta-model can be summarized to provide parameter estimates and confidence intervals, similar to those obtained via other methods. SHAP derives this explanatory meta-model from the outputs of a popular classification tree ensemble, XGBoost^22^. XGBoost is notably robust against multicollinearity^23^, automatically facilitates data curation^24^, and outperforms even the most state-of-the-art prediction methods for most types of structured data^25^.

Our aim was therefore to explore the utility of such a model in the context of prospective early warning systems. Herein, we report the application of SHAP to a large, high-resolution database consisting of variables from various domains – climate, environment, economic, sociodemographic, vector distribution, and host presence. The extraordinary WNV outbreak of 2018 in Europe was selected as the predictive objective due to the extreme degree to which it differed from prior years. For the early warning context, predictive accuracy is just as important as the accuracy of estimated feature effects^26^. Two important criteria for the latter are that parameter estimates are 1) reasonably invariant to model permutation and 2) broadly equivalent to those generated by other methods. Herein the former aim is addressed by developing several model permutations and comparing reported effects. The latter aim is then addressed by comparing and calibrating outputs of an established traditional model (Tran et al., 2014) against a comparably configured SHAP model.

## Results

### The 2018 WNV outbreak season was extraordinary in many ways

The number of regions reporting cases as well as the proportion of regions affected per country was substantially higher than in previous years (*Figure 1a*). Beyond that, many regions affected in 2018 had a history of WNV outbreak, but emergence has been overall sporadic (*Figure 1b*). Overall, observed WNV range was substantially higher in 2018 compared to control (134 regions vs 44.9 mean regions during the 2010-2016 training period; see *Figure 1c*), with 25.4% previously naive to WNV.

### Overall model performance consistently around 96% (AUC)

For the the four primary models (Q1, Q2, Q3, and Q4), the probability of correct out-of-sample identification of a WNV outbreak region was statistically identical (i.e., consistent) regardless of model specification: 96.2% average, with mutually overlapping 95% confidence intervals. Refer to *Table 1*. This degree of performance is particularly notable given the substantial proportion of previously naive regions affected in 2018 and overall sporadic nature of occurrence during the training period (2010-2016). Refer to *Figure 1*.

**Table 1.** Table of performance (AUC) metrics for each of the four models. AUC is the probability that a randomly selected positive case will be assigned a higher probability than a randomly selected negative case. It indicates how well the model discriminates between positive and negative classes irrespective of classification threshold. Given the overlapping confidence intervals, overall discriminatory performance is therefore statistically identical regardless of empirical content.

| Model Performance | AUC (95% CI) |  |
| --- | --- | --- |
|  | Train (2010-2016) | Test (2018) |
| Q1 | 100% (*) | 95.3% (93.5 - 97.1%) |
| Q2 | 100% (*) | 96.3% (94.5 - 98.1%) |
| Q3 | 100% (*) | 96.6% (94.9 - 98.3%) |
| Q4 | 100% (*) | 96.2% (94.5 - 97.9%) |

### Predictive performance is highly sensitive to threshold assumptions

Despite overall high model performance, threshold choice was found to severely impact actual predictive performance. Both heuristic optimization methods, Youden’s index and the AUC maximization criteria, consistently produced predictions that were overall more accurate than the SHAP estimation. This remained true across all model configurations. However, this accuracy comes at the cost of predictive specificity: about one out of every ten true negatives were falsely labeled positive. This is noteworthy given that 92.6% of the n=1811 NUTS3 regions in this same are true negative. SHAP consistently maintained a far higher level of specificity – less than one out of one-hundred negative predictions were false.

**Table 2a,b,c.** Table of predictive performance for each of the four models and three threshold criteria. Predictive performance is threshold-dependent. Sensitivity, also referred to as true positive rate, measures the probability of detecting event occurrence. Specificity, also referred to true negative rate, measures the probability of correctly identifying non-occurrence. Specificity is consistently high for the empirical, SHAP-optimized model. Heuristic criteria favor higher sensitivity, the result of which is a high rate of “false alarms”. Sensitivity for he SHAP-optimized model improves as empirical data is added to the model. The heuristic threshold criteria are insensitive in this respect.

| Youden | Threshold | Specificity | Sensivity | Bal. Acc. |
| --- | --- | --- | --- | --- |
| Q1 | 0.01% | 91.5% | 86.5% | 89.0% |
| Q2 | 0.12% | 95.6% | 87.2% | 91.4% |
| Q3 | 0.04% | 90.2% | 93.6% | 91.9% |
| Q4 | 0.03% | 89.2% | 94.3% | 91.7% |

| AUCmax | Threshold | Specificity | Sensivity | Bal. Acc. |
| --- | --- | --- | --- | --- |
| Q1 | 0.01% | 89.3% | 88.7% | 89.0% |
| Q2 | 0.06% | 91.3% | 90.8% | 91.0% |
| Q3 | 0.04% | 90.2% | 93.6% | 91.9% |
| Q4 | 0.03% | 89.2% | 94.3% | 91.7% |

| SHAP | Threshold | Specificity | Sensivity | Bal. Acc. |
| --- | --- | --- | --- | --- |
| Q1 | 0.74% | 99.9% | 26.2% | 63.1% |
| Q2 | 0.67% | 99.0% | 61.7% | 80.3% |
| Q3 | 0.63% | 99.3% | 54.6% | 77.0% |
| Q4 | 0.61% | 99.3% | 53.9% | 76.6% |

Higher sensitivity is preferred in cases where the underlying model or test is mechanistically robust: i. e., cases where classification is not likely to be spurious. A qualitative assessment of the predictive maps generated by each of these methods for each of these models [*Figure S1*] confirms. SHAP is consistently conservative and the quality of prediction improves as mechanistically relevant data is added. As described in the *Methods* section, SHAP predictions – while tied to the XGBoost probabilistic outputs – are associated with a deductively imputed experimental substructure. SHAP only reports positive indication when sufficient data exists to support (refer to *Figure S1, Q1*). No such considerations are included when heuristic methods are employed to compute thresholds.

### SHAP feature effect estimates are consistent with original XGBoost models

SHAP produces a transformed feature effect matrix. Feature importance as determined by XGBoost (“gain”) is therefore not necessarily the same as that determined by SHAP. *Figure 3* demonstrates the degree to which the two models diverge from one another with respect to each of the four model configurations. Note that the only substantial deviation appears in the top left corner: the autoregressive feature indicative of a local history of outbreak. Note that SHAP apparently reassigned importance away from this spatiotemporally inconsistent predictor towards others that are more consistently deterministic as deduced by SHAP.

**Figure 3.**
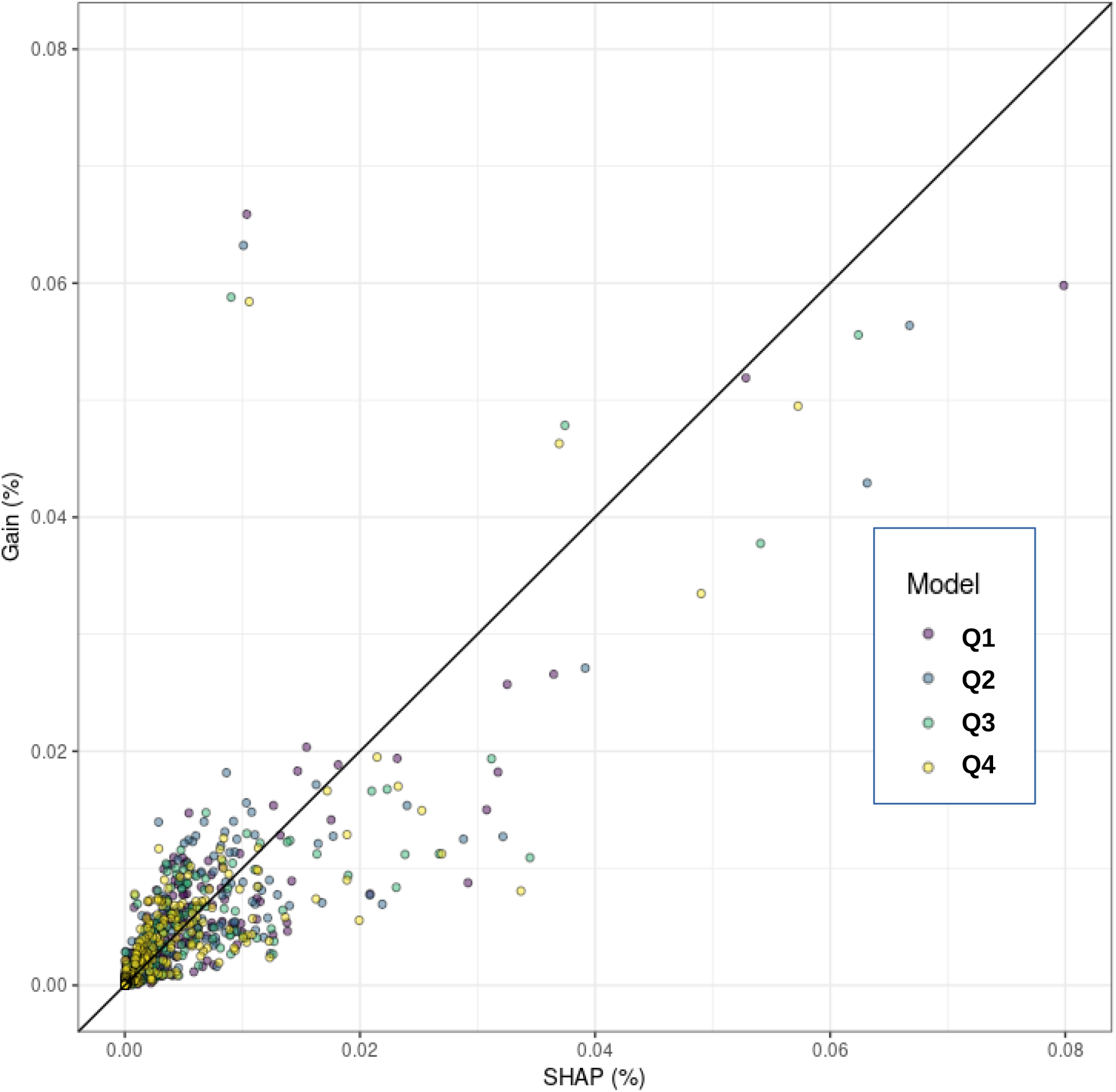
Feature-wise comparison of marginal contribution metrics: XGBoost vs SHAP. Feature importance, as empirically determined by SHAP, is largely consistent with that determined based solely on marginal contribution with respect to model output. The only notable difference is regarding the autoregressive feature. SHAP correctly identifies this feature as having substantially less probative power with respect to local prediction of outbreak risk; predictive weight is therefore reassigned to features with higher degrees of empirical relevance on a case-wise basis.

### SHAP model effects are consistent irrespective of model configuration

The SHAP effect matrix can be summarized to produce feature effect estimates. *Figure 4* shows aggregate effect estimates for regions with positive outbreak risk indication. The top features across models are vapor pressure, the autoregressive past year’s feature, maximum temperature, wind speed, and local GNP. Moreover, when aggregated across quarters, the effect of these features are broadly consistent across model configurations. Model Q1 is the notable exception (see *Discussion*).

**Figure 4.**
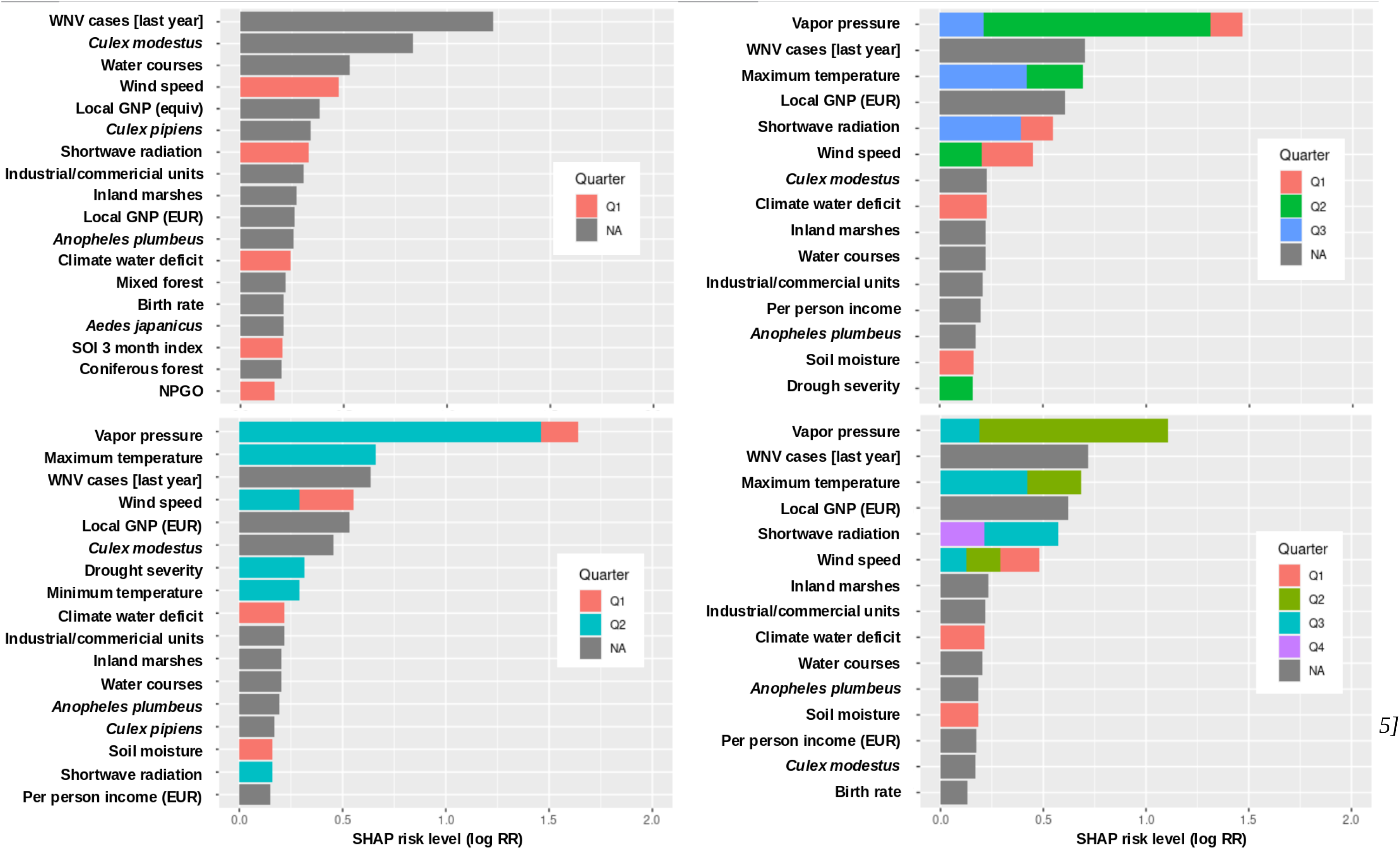
Top features and aggregate effect estimates for each model. SHAP deductively imputes feature effects based on observed local impact with respect to outbreak risk. This method of assignment is does not allow for spurious assignment of predictive weight. Features that demonstrate similar impact interactive effects will consequently be assigned effect scores that are consistent: this is part of the original specification of the SHAP heuristic proposed by Shapely in 1951. The SHAP specification also requires group effects to be equally valid to individual effects. Consequently, in cases where model specification consist of sequentially added data, that differs only with respect to temporal scope, features that are mechanistically identical will be assigned equivalent effect scores in the aggregate. Climatic effects as determined by SHAP are therefore consistent when aggregated across quarters. This consistency, along with presumed accuracy of predicted effect, improves as empirically relevant data is added to the model.

### Spatiotemporal and time-varying covariates improve performance, superficially

Features that capture spatiotemporal covariation perform well in isolation (AUC of 89% – 92%; *Figure S2*), as do time-varying bioclimatic features (AUC of 95% and 97% resp, logit and XGBoost; *Figure S3*). And in combination, these features deliver performance that is statistically equivalent to our fully-featured models (AUC of 96.2% and 96.7% resp., logit and XGBoost; *Figure S4*). And when added to our original early warning model (H1), the result is a model with superior predictive performance and exceptional discriminatory power (AUC 98.1%; *Figure S5*). Feature effect variability is demonstrably locale-dependent, with substantial bimodality of effect (*Figure 5a*) particularly with respect to temperature and temperature-dependent features (i.e, vapor pressure). It is important to note that covariates representing spatiotemporal covariation and time-varying bioclimatic features are user-defined. These can therefore be prospectively redefined and manipulated for the purpose of scenario-based forecasts. As previously noted, these covariates are overly sensitive in isolation and therefore produce empirically unreliable predictions. Note that the estimates produced by SHAP continue to be generally invariant to model permutation, even in this context. As shown in *Figure 5b*, this oversensitivity is then moderated by the effects of the retrospective climatic data as well as whatever structural, time-invariant factors have been found to be associated with a reduction in risk.

**Figure 5.**
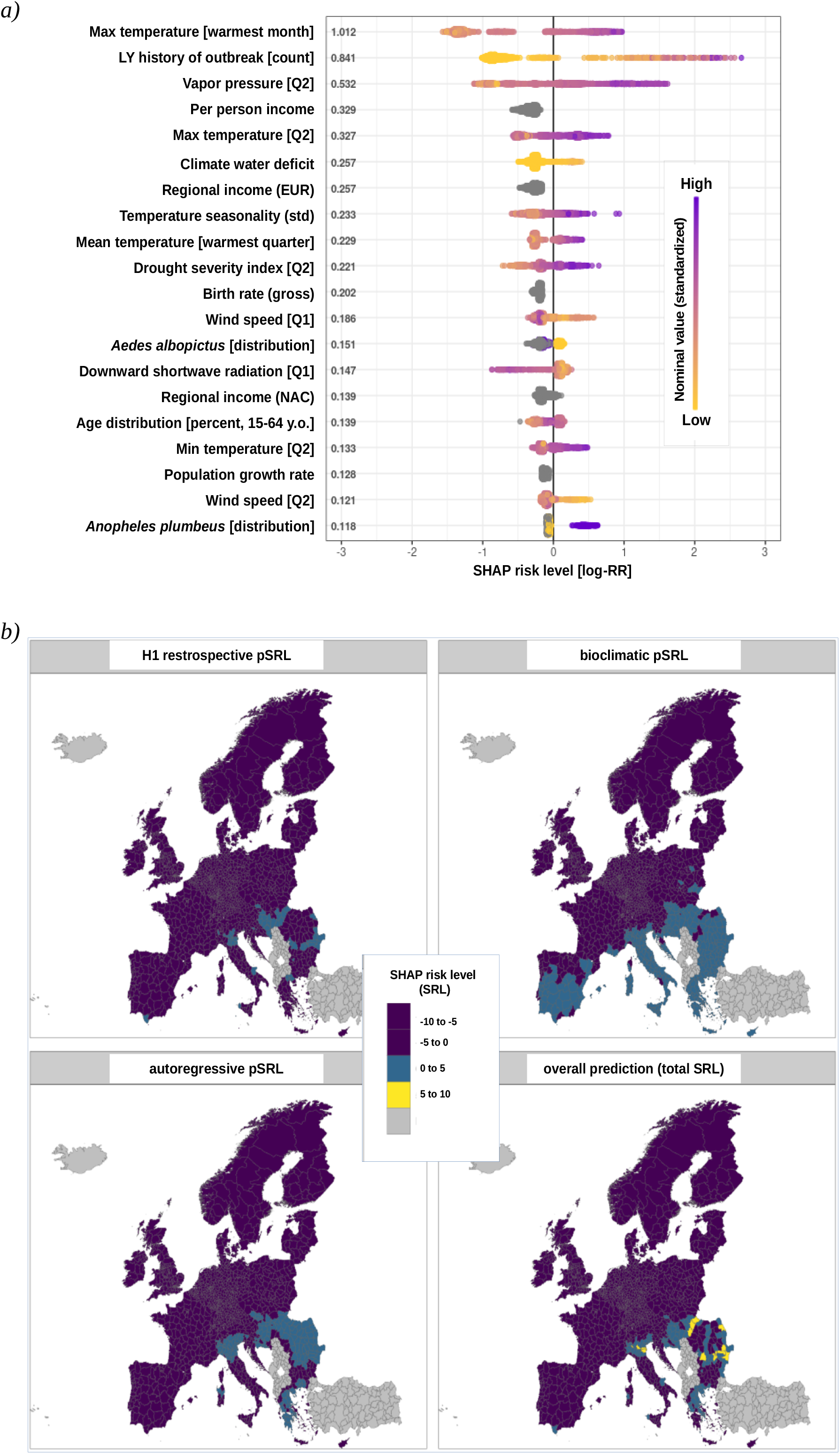
Local effect variability confirmed by density plot and geospatial map. a) Density plot that visualizes the effect distributions for each case on a feature-wise basis b) Additive maps demonstrating the independent effect of the various features sets as well as how they combine to produce the final outbreak prediction. **Local effect variability confirmed by density plot and geospatial map** Figure 5a demonstrates the degree of geospatial variability and effect bimodality revealed by SHAP. This finding is consistent with the research literature that notes substantial variability in reported feature effects (see discussion). SHAP is specified to allow for direct model decomposition: this, in turn, allows for examination of the independent, aggregate effect of any set of features as well as observation of their additive effects with respect to overall predicted risk *(Figure 5b)*.

As previously shown, feature effect estimates can be derived by aggregating regions with positive outbreak indication. However, no method exists to generate estimates that would compare to those produced by traditional methods. That said, ranked feature importance along with an analysis of effect distribution provides directional insight appropriate for comparison to the literature. The following table presents the top 10 most important features for prediction, as determined by SHAP.

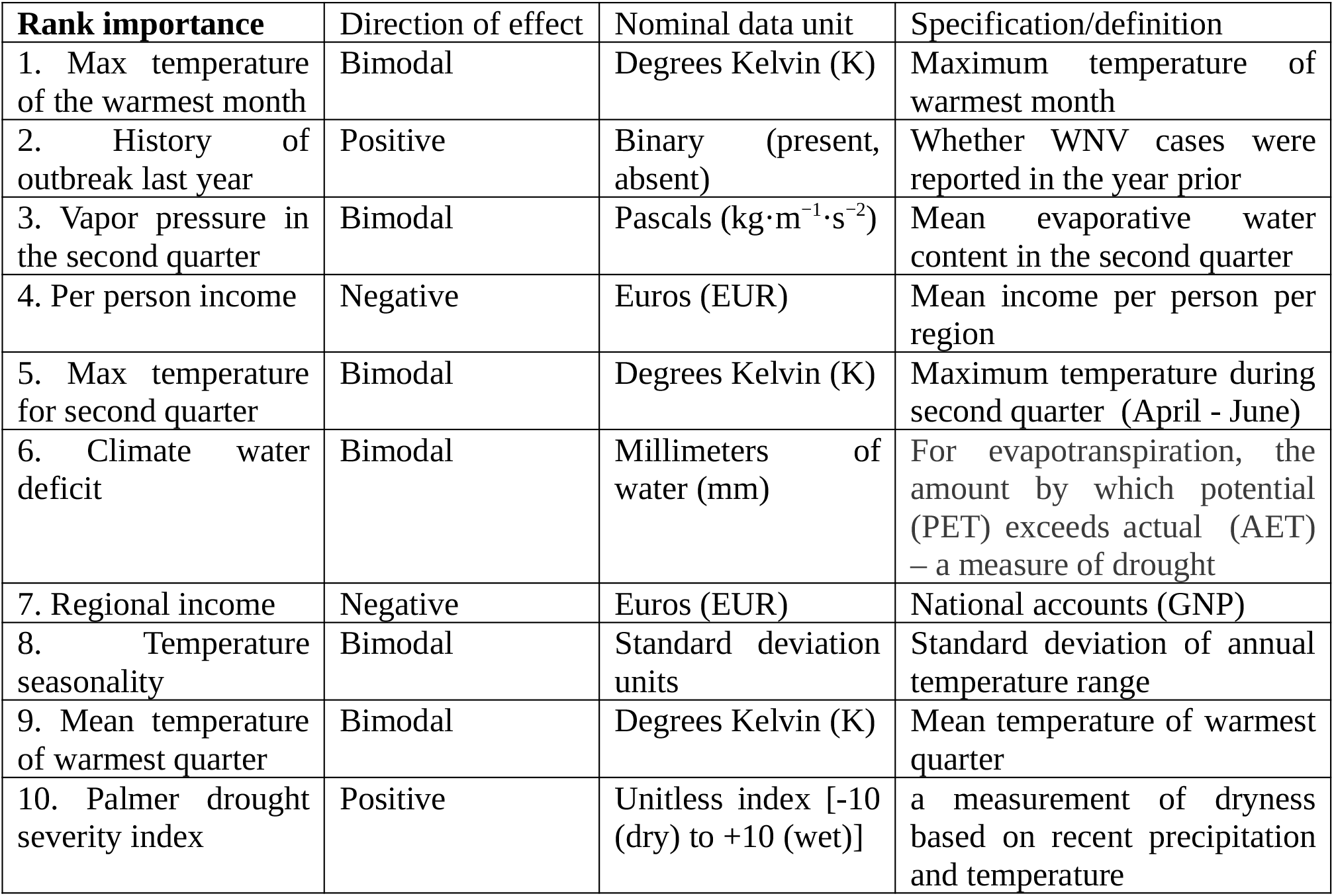

## Discussion

Predictive modeling efforts thus far have been confounded by several unavoidable realities. WNV is particularly difficult due to the complexity associated with an enzootic transmission cycle involving undetermined reservoir hosts and markedly demonstrated vectorial plasticity. As demonstrated in this study, models with limited or empirically imprecise data can therefore produce implausible predictive results. This study demonstrates the degree to which an AI-driven feature effect estimation engine (SHapely Additive eXplanations; SHAP) to mitigate such concerns. SHAP generates empirically-precise models for each individual case and sets predictive thresholds accordingly. As demonstrated by the present study, specificity is therefore optimized at the expense of sensitivity: despite having the same probabilistic base as the XGBoost model, classification thresholds are set such that predictions with less or lower-quality statistical support are suppressed. To our knowledge, no other method exists for empirically determining classification thresholds.

The determinants driving the emergence and spread of WNV are many and diverse. As such traditional modeling option cannot reasonably capture the full spectrum of variability. This is due to many causes, but mostly reflect the inability of traditional linear models to adequately handle multicolinearity and bimodality or localization of effect. Such limitations are inherent to the linear estimation process and are therefore not able to be overcome by any degree of methodological diligence. As demonstrated in the present work, XGBoost has the notable advantage of being immune to such limitations and therefore is able to consider feature set sizes that number in the hundreds. And when SHAP is applied, the underlyng variability represented by the data is exposed and therefore able to be exploited to deliver empirically robust predictions, with a high level of specificity. Up to now, “black box” machine learning has suffered from poor generalizeability due to a lack of empirical grounding. A brief literature review illustrates the complexities involved.

### Determinants of West Nile Virus in the Literature

Climate change and variability has now been well-established as a driver of the spread and severity of infectious disease outbreaks^27,28^across the globe. Many factors are known to contribute to this association and research is ongoing to uncover and explore more. Climate has long been established as being a driver of biotic and abiotic factors that contribute to disease^29,30^, as has changing patterns to and increases in human mobility^31,32^. Changing climate has also been associated with changes to the migratory patterns of disease-carrying birds^33^ and has been suggested to contribute to the dispersal of disease-carrying mosquito vectors^34^. And such trends are projected to accelerate^35^.

The severity and spread of infectious disease across endemic regions and beyond has been furthermore linked to regular, long-term climate variability. For example, variability driven by ENSO has long been established to be a clear driver of dengue and malaria in the sub/tropical^36^ and South Pacific^37^ regions. And in Europe, the equivalent NAO cycle has similarly been associated with an entire host of diseases including infectious mononucleosis, salmonellosis, erysipelas, toxoplasmosis (positive) as well as hepatitis A, scarlet fever, leptospirosis, shigellosis (negative)^38^.

There are many mechanisms according to which climate contributes to the enhancement and spread of disease such as West Nile virus^39^. One key underlying driver is the ongoing change to the habitats and behavior of both hosts and vectors^40,41^. This is especially notable in the case of mosquito borne disease^42^, particularly those capable of being carried long-distance by regular host migration. Changes to conditions in the northern latitudes of both Europe and America have increased the range and susceptibility of the mosquitos responsible for transmission^43,44^. The invasion and establishment of non-native, but disease-competent mosquito species has likewise also been associated with improved habitat suitability due to weather that is warmer and wetter^45,46^.

Much research attention has been focused on more precisely understanding these dynamics. There are two broad categories of quantitative models applied for this purpose: statistical and process-oriented. Statistical models focus mainly on the analysis of time-series and patterns associated with disease outbreak data. The purpose of such models is to predict occurrence, and perhaps also severity, absent mechanistic justification^19^. They are generally regarded as being more statistically precise, but suffer from overfitting and problematic generalizability^47^. Such models are commonly used for development of early warning systems and for intervention (resource) planning^48^. The XGBoost model presently presented would fall under this category absent the application of SHAP.

Process-oriented, mechanistic, models set out to more precisely quantify and characterize the relationship between known and suspected causal determinants and other factors associated with observed outbreaks of disease. Such mechanistic models attempt to extend theory and practical knowledge to the problem of predicting and controlling the factors driving the manifestation and spread of disease^49^. Questions that might be addressed within the context of such models include—a) association between vector abundance and disease manifestation^50^, b) climate- and environment-associated biotic factors that influence “a”^27,51^, c) population vulnerability factors that influence “b”^52^, d) human behavioral and mobility patterns that influence “c”^32,53^, and e) the effects of various control/clinical interventions on all of the above^54^.

Beyond that, climate change scenarios based on long-term climate change projections have been used to estimate the future spread of arbovirus vectors and associated outbreak risks. Such studies have relied upon presumed associations between determinants of arbovirus outbreak risk and climate change, both with respect for changes to vector habitat suitability^55^ and in association with socioeconomic development and consequent anthropogenic changes to the environment^56^. These presumed associations have, however, never been firmly quantified and are simply educated guesses at present. Absent a suitably robust analytical method and appropriate data, it is exceedingly difficult to develop an empirically sound basis for long-term projections. Up to now, progress in this arena has been stymied by the frequent observation that, for many climate-related variables, causal associations are time-dependent and appear to change depending on time-lag and seasonality^19^. In addition, the impact of undercounts is an ongoing concern^57^, both with respect to predicting the timing and severity of disease outbreaks and interpretation of the underlying mechanisms inferred. The work presented herein offers the first concrete steps towards resolution of this matter. As demonstrated, the effects reported by SHAP are relatively robust to model permutation and can therefore be presumed reliable long term. The degree to which these effect estimates can be used for long-term projections is a matter that will require further study.

Beyond such higher-level drivers of outbreak risk, day-to-day human behavior is the ultimate driver of infection and manifestation of disease^2^. Sociodemographic statistics such as age, gender, and occupation serve as useful proxies. Higher level behavior associated with economic statistics, such as human mobility and increased trade, has also been associated with increased outbreak risk^58^ – a finding supported by the present work. This is especially important in the context of arboviruses such as Japanese encephalitis, zika, dengue, utsutu, and—yes—West Nile virus for which economic activity has been linked to the introduction of both disease^53^ and disease-competent vectors in Europe^59^. Up to now, however, individual-level features have been difficult to include in modeling efforts. The presently reported model demonstrates the probative value of including such features and our hope is that other will continue to incorporate such features, especially in light of the high potential long-term disease burdens associated with even asymptomatic cases^60^. This is an oversight that must urgently be addressed—especially in the context of accelerating and spreading arbovirus outbreak risk associated with variability in climate and anthropogenic change.

### Conclusions

Any given event can be attributed to any given number of actors or supervening features. And many statistical, inferential methods exist to estimate such effects. Each of these methods has its pros and cons, and new ones are constantly on the horizon. Up till recently, however, derivation of AI-derived effect estimations has been computationally prohibitive. The present work demonstrated a novel implementation of AI applied to machine learning to derive high-quality effect estimations. We show that that these effect estimations are relatively invariant to model permutation and that aggregate effect estimations remain consistent even when features are arbitrarily disaggregated. We furthermore show that when applied to the context of geospatial disease prediction and early warning, the Shapley criteria enforces a strict empirical robustness on the output geospatial predictions. We compare to more traditional models following traditional criteria and show that the lack of such criteria results in spurious, mechanistically infeasible, predictions as model sensitivity is increased. Up to now, the machine learning world has effectively ignored this aspect – pursuing predictive power above all other considerations. This has unfortunately produced the paradoxical effect whereby high-performance ML-based models cannot be reliably deployed for long-term prediction and generalized forecasting. The method presented promises to change this for the better and open the door to new world of epidemiological precision. These models were validated against the extraordinary 2018 outbreak season to demonstrate the degree to which such methods can yield high-quality predictions even when the environment substantially changes. This is especially important given the context of climate-driven changes to infectious disease potential.

## Data Availability

Data available upon request.

## Acknowledgements

A.A.G. received funding from the European Center for Disease Control and Prevention (contract no ECDC.9504).

## Author contributions

The authors worked collaboratively and contributed equally to the article.

---

**Figure S1.**
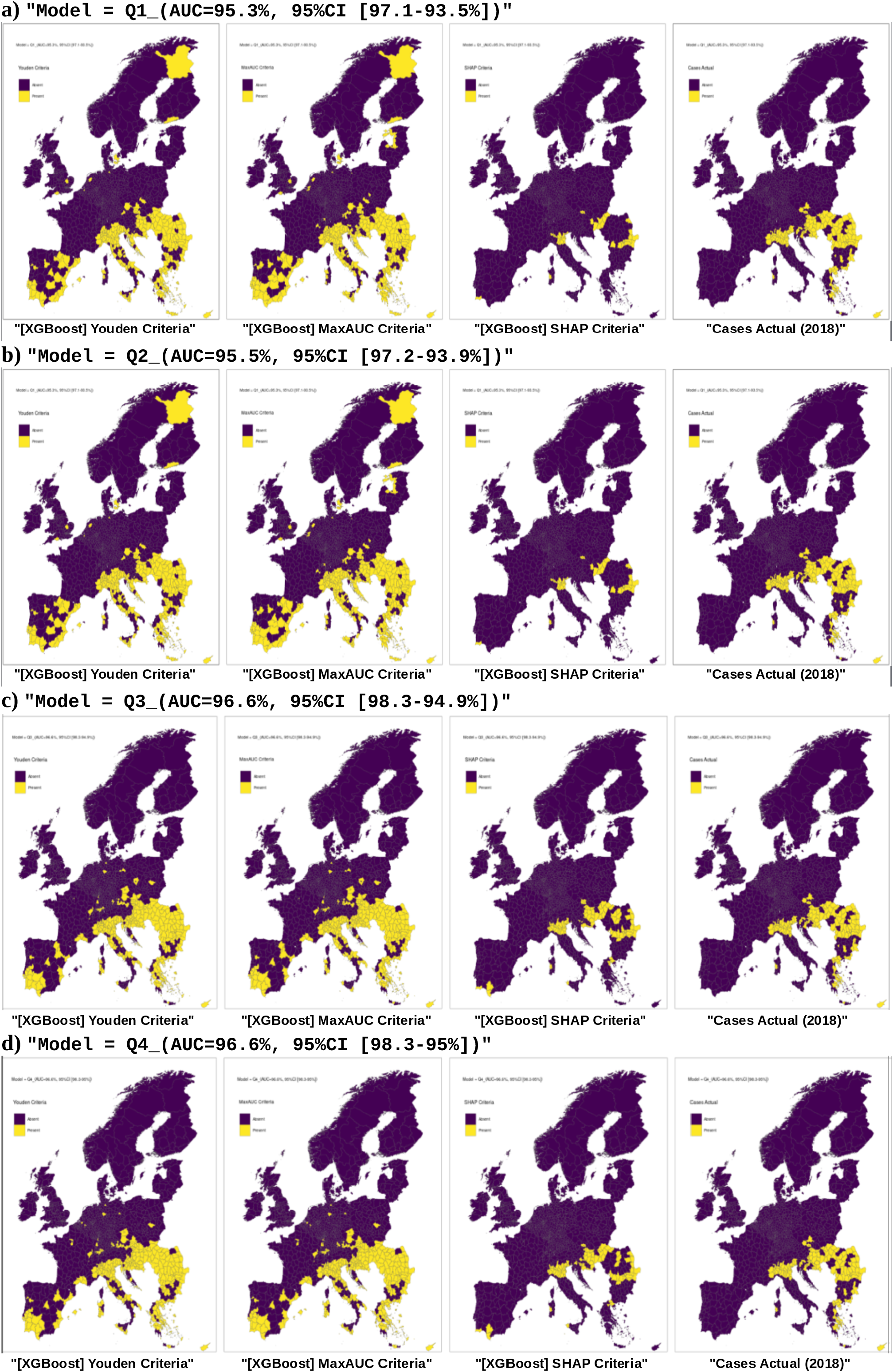
SHAP-based predictions appear to converge as data is added. SHAP predictions are based on empirical support and therefore conservative compared to those derived via conventional “top-down” optimization methods. As empirically relevant data is added to the model, the SHAP-generated predictive map converges with the actual case-reported map. However, this is not the case for the predictions derived via the conventional, empirically-blind methods. These models are consistently over-sensitive and show no tendency to converge to the actual, on-the-ground solution.

**Figure S2.**
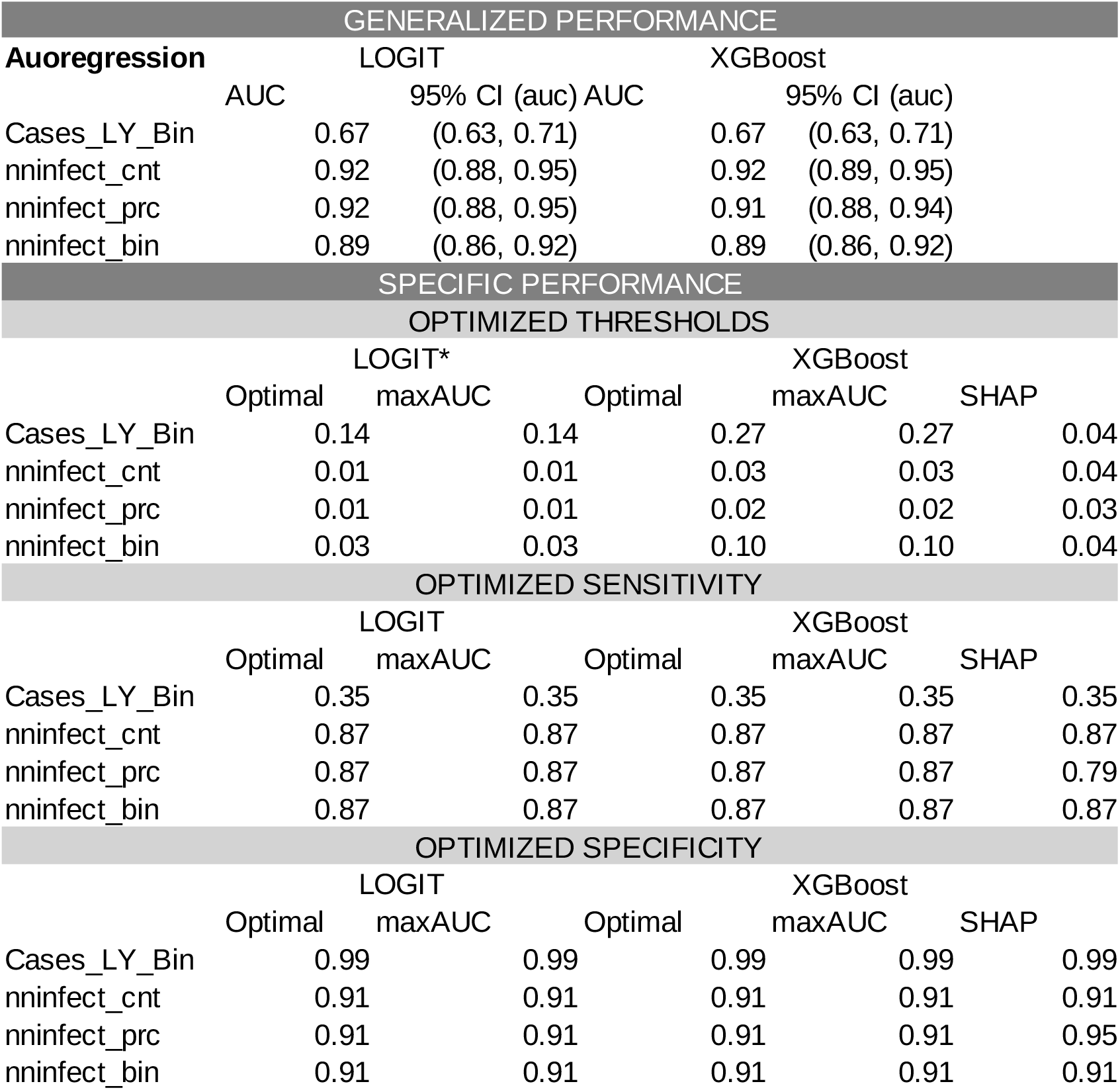
Autoregressive geospatial covariation alone performs reasonably well. Autoregressive features derived from Tran et al. (2014) incorporating geospatial covariation were tested: 1) number of neighboring districts (self-inclusive) with reported outbreak in prior year [nninfect_cnt (A1)], 2) percentage of neighboring districts (self-inclusive) with reported outbreak in prior year [nninfect_prc (A2)], and 3) outbreak reported in same or neighboring district in prior year [nninfect_bin (A3)]. For benchmark purposes the purely autoregressive covariate featured in our models presented thus far, ‘outbreak reported in same district in prior year’ [Case_LY_Bin (A0)], was also evaluated. In all cases, the covariates derived from the Tran et al (2014) model performed well – even exceeding the performance originally reported within Tran et al (2014). The underperformance demonstrated by the purely autoregressive feature confirms that geospatial covariation is indeed statistically relevant; however, predictive performance is limited absent empirical support. As shown in our primary models, SHAP effectively provides for such support given sufficient quantity and quality of data.

**Figure S3.**
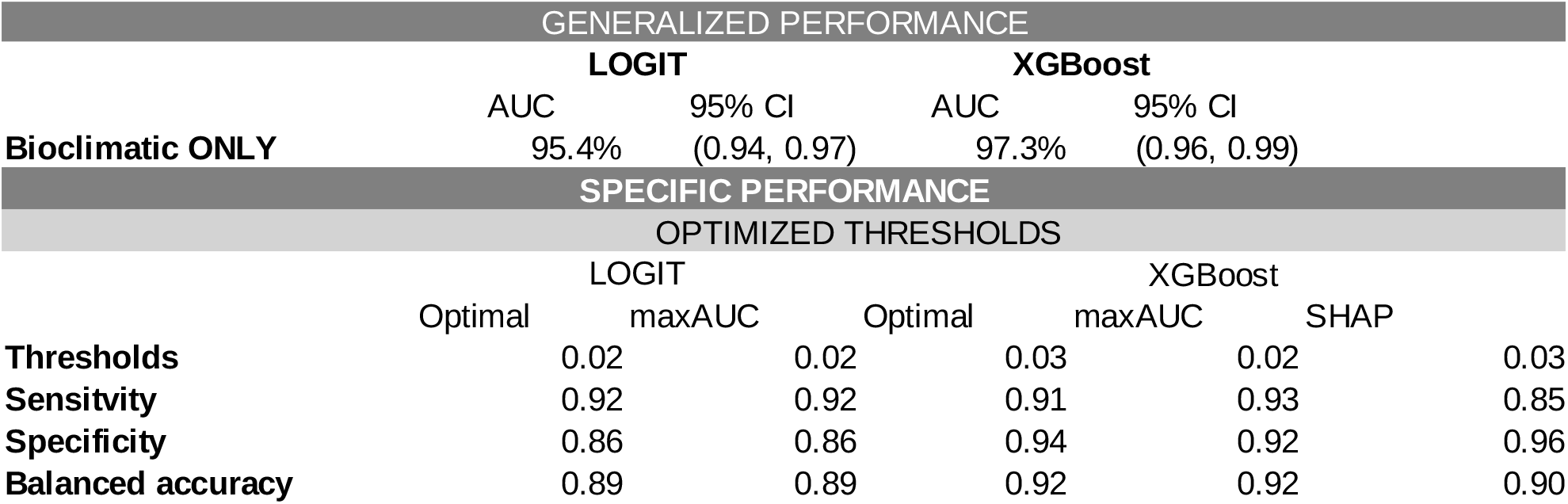
Time-varying bioclimatic features alone also perform reasonably well. Bioclimatic features are calculated based on broad temporal trends are meant to be relatively stable year-to-year. The unpublished exploratory model, from which the present study is derived, reports these features to be of high importance for prediction. However, the bioclimatic features require an entire years data to calculate and are therefore unsuitable for the early warning use case. We nevertheless generated and evaluated a model featuring only these bioclimatic features (B1). Retrospective performance using only these bioclimatic features was found to be excellent. As demonstrated in the previously cited exploratory model, these features serve remarkably well for retrospective explanation. Indeed, the advantage provided by XGBoost and SHAP with respect to empirical validity is minimized in this model. Given that these features are user-defined, one could conceivably use them as basis for prospective, scenario-based long-term forecasts – provided said scenarios are empirically justified.

**Figure S4.**
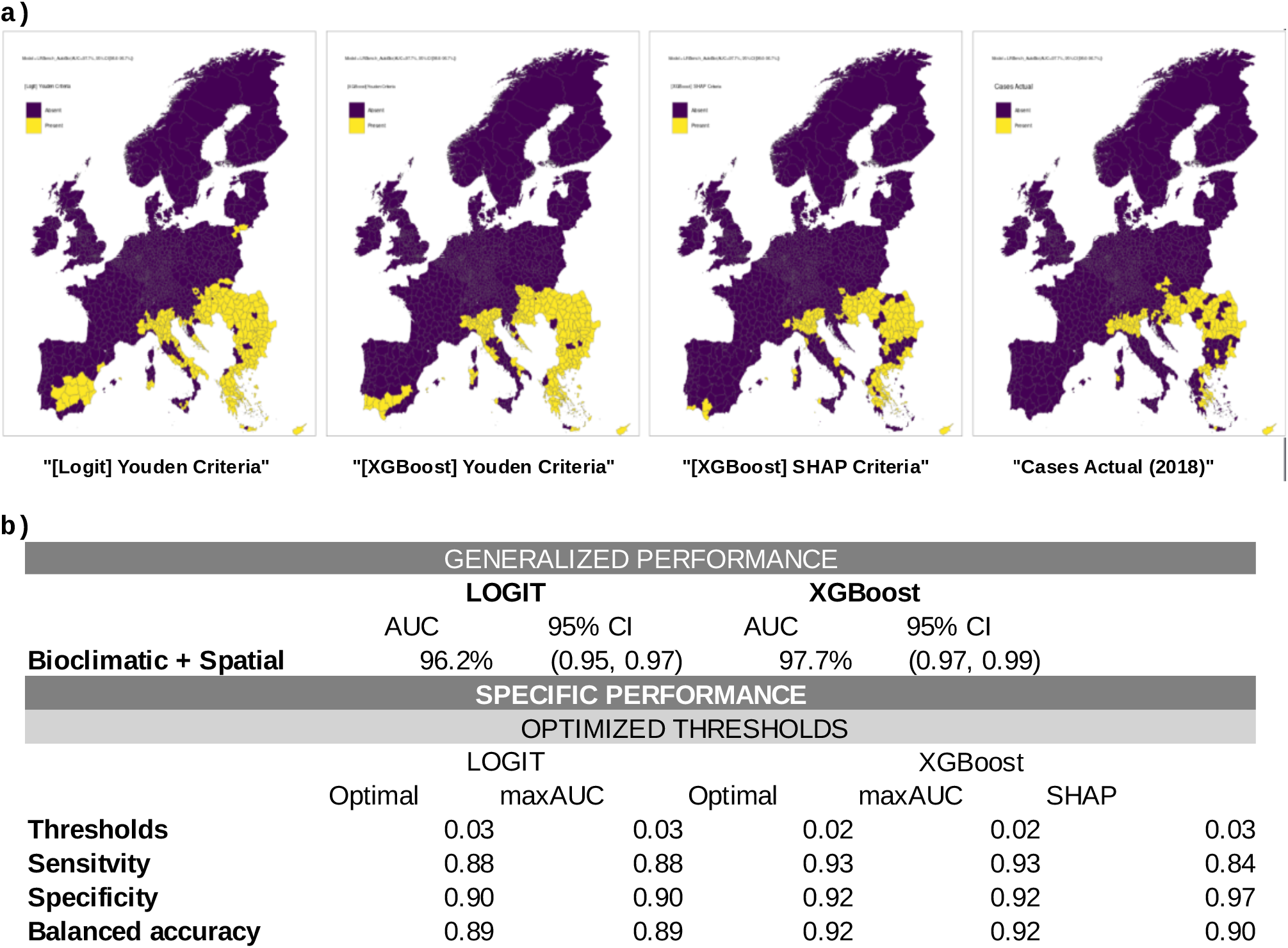
Combination of generic features provides little benefit. A diagnostic model combining the spatiotemporal and time-varying, bioclimatic covariates (B1+) showed negligible improvement. Given how the bioclimatic variables are derived – 30 years regional averages with respect to various climatic indices – an autocorrelative effect is expected. In this simple model, no tangible benefit is observed in terms of model performance or predictive specificity – SHAP lacks sufficient interactive information to deduce differential effects. As we later observe, this however changes with the addition of a broader spectrum of empirical data.

**Figure S5.**
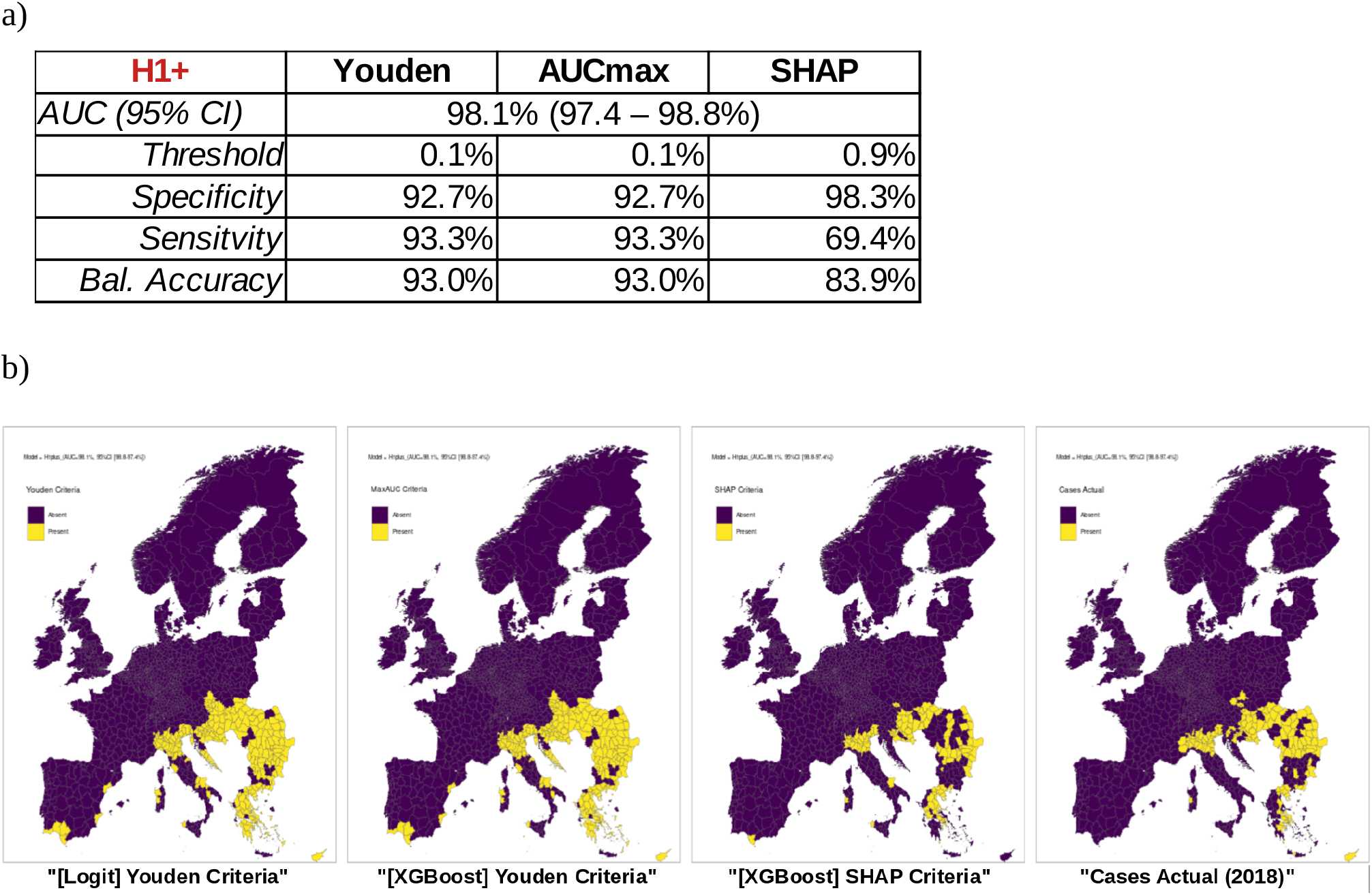
High-precision early warning and scenario-based forecasts are possible with SHAP. A final model incorporating the early-warning scope (H1), auotregressive geospatial covariation (A1), and time-varying bioclimatic variables (B1) delivered the performance approaching 99% AUC (97.4 – 98.8%). By combining restrospective data and prospective estimations based on prior years, this model allows for high-precision, same-year forecasting of West Nile virus emergence. Beyond this high out-of-the-box performance, this model more interestingly allows for scenario-based forecasting via respecification of the autoregressive and time-varying features. Additional work will be required to better qualify the practical scope of scenario-building and to quantify performance.

## References

1. Patel, H., Sander, B. & Nelder, M. P. Long-term sequelae of West Nile virus-related illness: a systematic review. Lancet Infect. Dis. 15, 951–959 (2015).

2. Bárdos, V. et al. Neutralizing Antibodies against some Neurotropic Viruses determined in Human Sera in Albania. J. Hyg. Epidemiol. Microbiol. Immunol. 3, 277–82 (1959).

3. De Filette, M., Ulbert, S., Diamond, M. S. & Sanders, N. N. Recent progress in West Nile virus diagnosis and vaccination. Vet. Res. 43, 16 (2012).

4. Guenno, B. L., Bougermouh, A., Azzam, T. & Bouakaz, R. West Nile: a deadly virus? The Lancet 348, 1315 (1996).

5. Zeller, H. G. & Schuffenecker, I. West Nile Virus: An Overview of Its Spread in Europe and the Mediterranean Basin in Contrast to Its Spread in the Americas. Eur. J. Clin. Microbiol. Infect. Dis. 23, 147–156 (2004).

6. Vittor, A. Y. et al. West Nile Virus-Induced Neurologic Sequelae—Relationship to Neurodegenerative Cascades and Dementias. Curr. Trop. Med. Rep. 7, 25–36 (2020).

7. Hernández-Triana, L. M. et al. Emergence of West Nile Virus Lineage 2 in Europe: A Review on the Introduction and Spread of a Mosquito-Borne Disease. Front. Public Health 2, (2014).

8. Camp, J. V. & Nowotny, N. The knowns and unknowns of West Nile virus in Europe: what did we learn from the 2018 outbreak? Expert Rev. Anti Infect. Ther. 18, 145–154 (2020).

9. Di Sabatino, D. et al. Epidemiology of West Nile Disease in Europe and in the Mediterranean Basin from 2009 to 2013. BioMed Research International vol. 2014 e907852 https://doi.org/10.1155/2014/907852 (2014).

10. Pandit, P. S. et al. Predicting wildlife reservoirs and global vulnerability to zoonotic Flaviviruses. Nat. Commun. 9, 5425 (2018).

11. Root, J. J. & Bosco-Lauth, A. M. West Nile Virus Associations in Wild Mammals: An Update. Viruses 11, 459 (2019).

12. Hubálek, Z. & Halouzka, J. West Nile fever--a reemerging mosquito-borne viral disease in Europe. Emerg. Infect. Dis. 5, 643–650 (1999).

13. Hadjichristodoulou, C. et al. West Nile Virus Seroprevalence in the Greek Population in 2013: A Nationwide Cross-Sectional Survey. PLoS ONE 10, (2015).

14. Nagy, A., Szöllősi, T., Takács, M., Magyar, N. & Barabás, É. West Nile Virus Seroprevalence Among Blood Donors in Hungary. Vector-Borne Zoonotic Dis. 19, 844–850 (2019).

15. Panayotova, E. et al. SEROPREVALENCE OF WEST NILE VIRUS IN BULGARIA, 2018. Probl. Infect. Parasit. Dis. 47, 15–17 (2019).

16. Keeling, M. J. & Rohani, P. Modeling Infectious Diseases in Humans and Animals. (Princeton University Press, 2011).

17. Chianese, A. et al. West Nile virus: an overview of current information. Transl. Med. Rep. 3, (2019).

18. Dodd, R. Y., Foster, G. A. & Stramer, S. L. Keeping Blood Transfusion Safe From West Nile Virus: American Red Cross Experience, 2003 to 2012. Transfus. Med. Rev. 29, 153–161 (2015).

19. Anesi, J. A. & Silveira, F. P. Arenaviruses and West Nile Virus in solid organ transplant recipients: Guidelines from the American Society of Transplantation Infectious Diseases Community of Practice. Clin. Transplant. 33, e13576 (2019).

20. Centers for Disease Control and Prevention (CDC). West Nile virus infection among turkey breeder farm workers--Wisconsin, 2002. MMWR Morb. Mortal. Wkly. Rep. 52, 1017–1019 (2003).

21. Centers for Disease Control and Prevention (CDC). Laboratory-acquired West Nile virus infections--United States, 2002. MMWR Morb. Mortal. Wkly. Rep. 51, 1133–1135 (2002).

22. Hayes, E. B. & O’Leary, D. R. West Nile Virus Infection: A Pediatric Perspective. Pediatrics 113, 1375–1381 (2004).

23. Pisani, G., Cristiano, K., Pupella, S. & Liumbruno, G. M. West Nile Virus in Europe and Safety of Blood Transfusion. Transfus. Med. Hemotherapy 43, 158–167 (2016).

24. Mrzljak, A. et al. West Nile Virus: An Emerging Threat in Transplant Population. Vector-Borne Zoonotic Dis. (2020) doi:10.1089/vbz.2019.2608.

25. Pachler, K. et al. Putative new West Nile virus lineage in Uranotaenia unguiculata mosquitoes, Austria, 2013. Emerg. Infect. Dis. 20, 2119 (2014).

26. Vázquez, A. et al. Putative new lineage of West Nile virus, Spain. Emerg. Infect. Dis. 16, 549 (2010).

27. Bondre, V. P., Jadi, R. S., Mishra, A. C., Yergolkar, P. N. & Arankalle, V. A. West Nile virus isolates from India: evidence for a distinct genetic lineage. J. Gen. Virol. 88, 875–884 (2007).

28. Shahhosseini, N. & Chinikar, S. Genetic evidence for circulation of Kunjin-related West Nile virus strain in Iran. J. Vector Borne Dis. 53, 384 (2016).

29. McMullen, A. R. et al. Molecular evolution of lineage 2 West Nile virus. J. Gen. Virol. 94, 318–325 (2013).

30. Ruiz, M. O., Tedesco, C., McTighe, T. J., Austin, C. & Kitron, U. Environmental and social determinants of human risk during a West Nile virus outbreak in the greater Chicago area, 2002. Int. J. Health Geogr. 3, 8 (2004).

31. Davis, J. K. et al. Improving the prediction of arbovirus outbreaks: A comparison of climate-driven models for West Nile virus in an endemic region of the United States. Acta Trop. 185, 242–250 (2018).

32. Barker, C. M. Models and Surveillance Systems to Detect and Predict West Nile Virus Outbreaks. J. Med. Entomol. 56, 1508–1515 (2019).

33. Potter, K. M., Koch, F. H., Oswalt, C. M. & Iannone, B. V. Data, data everywhere: detecting spatial patterns in fine-scale ecological information collected across a continent. Landsc. Ecol. 31, 67–84 (2016).

34. Georganos, S. et al. Very High Resolution Object-Based Land Use-Land Cover Urban Classification Using Extreme Gradient Boosting. IEEE Geosci. Remote Sens. Lett. 15, 607–611 (2018).

35. Lundberg, S. M. et al. From local explanations to global understanding with explainable AI for trees. Nat. Mach. Intell. 2, 56–67 (2020).

36. Lundberg, S. M., Erion, G. G. & Lee, S.-I. Consistent Individualized Feature Attribution for Tree Ensembles. ArXiv180203888 Cs Stat (2019).

37. Chen, T. & Guestrin, C. Xgboost: A scalable tree boosting system. in Proceedings of the 22nd acm sigkdd international conference on knowledge discovery and data mining 785–794 (2016).

38. Munkhdalai, L., Munkhdalai, T., Namsrai, O.-E., Lee, J. & Ryu, K. An Empirical Comparison of Machine-Learning Methods on Bank Client Credit Assessments. Sustainability 11, 699 (2019).

39. He, X. et al. Practical Lessons from Predicting Clicks on Ads at Facebook. in Proceedings of 20th ACM SIGKDD Conference on Knowledge Discovery and Data Mining – ADKDD’14 1–9 (ACM Press, 2014). doi:10.1145/2648584.2648589.

40. Nielsen, D. Tree boosting with xgboost-why does xgboost win” every” machine learning competition? (NTNU, 2016).

41. Hoover, K. C. & Barker, C. M. West Nile virus, climate change, and circumpolar vulnerability. WIREs Clim. Change 7, 283–300 (2016).

42. Medlock, J. M., Snow, K. R. & Leach, S. Potential transmission of West Nile virus in the British Isles: an ecological review of candidate mosquito bridge vectors. Med. Vet. Entomol. 19, 2–21 (2005).

43. Anopheles plumbeus – Factsheet for experts. European Centre for Disease Prevention and Control https://www.ecdc.europa.eu/en/disease-vectors/facts/mosquito-factsheets/anopheles-plumbeus.

44. Jelinski, D. E. & Wu, J. The modifiable areal unit problem and implications for landscape ecology. Landsc. Ecol. 11, 129–140 (1996).

45. Cohen, J. M. et al. Spatial scale modulates the strength of ecological processes driving disease distributions. Proc. Natl. Acad. Sci. 113, E3359–E3364 (2016).

46. Jaenson, T. G. T., Lokki, J. & Saura, A. Anopheles (Diptera: Culicidae) and Malaria in Northern Europe, with Special Reference to Sweden. J. Med. Entomol. 23, 68–75 (1986).

47. Najdenski, H. et al. Migratory birds along the Mediterranean – Black Sea Flyway as carriers of zoonotic pathogens. Can. J. Microbiol. 64, 915–924 (2018).

48. Michel, F. et al. Evidence for West Nile Virus and Usutu Virus Infections in Wild and Resident Birds in Germany, 2017 and 2018. Viruses 11, 674 (2019).

49. Semenza, J. C. et al. Climate change projections of West Nile virus infections in Europe: implications for blood safety practices. Environ. Health 15, S28 (2016).

50. Semenza, J. C. et al. Climate change projections of West Nile virus infections in Europe: implications for blood safety practices. Environ. Health 15, S28 (2016).

51. Lindgren, E., Andersson, Y., Suk, J. E., Sudre, B. & Semenza, J. C. Monitoring EU Emerging Infectious Disease Risk Due to Climate Change. Science 336, 418–419 (2012).

52. Lillepold, K., Rocklöv, J., Liu-Helmersson, J., Sewe, M. & Semenza, J. C. More arboviral disease outbreaks in continental Europe due to the warming climate? J. Travel Med. 26, (2019).

53. Shapley, L. S. Notes on the n-Person Game—II: The Value of an n-Person Game. (1951).

54. Cox, D. R. Regression models and life-tables. J. R. Stat. Soc. Ser. B Methodol. 34, 187–202 (1972).

55. Capelli, G. et al. First report in italy of the exotic mosquito species Aedes (Finlaya) koreicus, a potential vector of arboviruses and filariae. Parasit. Vectors 4, 188 (2011).

56. Komar, N. West Nile virus: epidemiology and ecology in North America. Adv. Virus Res. 61, 185–234 (2003).

57. Mancini. Mosquito species involved in the circulation of West Nile and Usutu viruses in Italy. Vet. Ital. 53, 97–110 (2017).

58. Hubálek, Z. European Experience with the West Nile Virus Ecology and Epidemiology: Could It Be Relevant for the New World? Viral Immunol. 13, 415–426 (2000).

59. Komar, N. et al. Experimental infection of North American birds with the New York 1999 strain of West Nile virus. Emerg. Infect. Dis. 9, 311 (2003).

60. Schönenberger, A. C. Host preferences of host-seeking and blood-fed Swiss mosquitoes. (2015) doi:10.5167/UZH-120144.

61. Martinet, J.-P., Ferté, H., Failloux, A.-B., Schaffner, F. & Depaquit, J. Mosquitoes of North-Western Europe as Potential Vectors of Arboviruses: A Review. Viruses 11, 1059 (2019).

62. Cohen, J. Statistical power analysis. Curr. Dir. Psychol. Sci. 1, 98–101 (1992).

